# Factors associated with acceptance of a digital contact tracing application for COVID-19 in the Japanese working-age population

**DOI:** 10.1101/2021.10.28.21265601

**Authors:** Tomohiro Ishimaru, Koki Ibayashi, Masako Nagata, Seiichiro Tateishi, Ayako Hino, Mayumi Tsuji, Hajime Ando, Keiji Muramatsu, Yoshihisa Fujino, for the CORoNaWork Project

**Author notes:** **Corresponding Author:** Tomohiro Ishimaru, MD, MPH, PhD, Department of Environmental Epidemiology, Institute of Industrial Ecological Sciences, University of Occupational and Environmental Health, Japan, 1-1 Iseigaoka, Yahata-nishi-ku, Kitakyushu, Fukuoka 807-8555, Japan. **Ethical Considerations & Disclosures** This study was approved by the Ethics Committee of the University of Occupational and Environmental Health, Japan (R2-079).

## Abstract

**Objective:** This study aimed to determine factors associated with acceptance of a Digital Contact Tracing (DCT) app for Coronavirus Disease 2019 (COVID-19) in the Japanese working-age population.

**Methods:** A cross-sectional study was performed for 27,036 full-time workers registered with an internet survey company during December 2020 in Japan.

**Results:** The rate of downloading the DCT app was 25.1%. The DCT app was more likely to be accepted by people with married status, university graduation or above, higher income, and occupations involving desk work. Fear of COVID-19 transmission, wearing a mask, using hand disinfection, willingness to be vaccinated against COVID-19, and presence of an acquaintance infected with COVID-19 were also associated with a greater likelihood of adopting the app.

**Conclusions:** The present findings have important implications for widespread adoption of DCT apps in working-age populations in Japan and elsewhere.

## Introduction

Digital Contact Tracing (DCT) has been used in many countries in response to the coronavirus disease 2019 (COVID-19) pandemic. DCT apps implemented in mobile devices (e.g. smartphones) can be employed to detect close contact with infected individuals and prevent the spread of COVID-19.^1^ Although the method of contact tracing has been used for a long time, traditional paper-based surveys are complicated and time-consuming, and are affected by loss of questionnaires and huge amounts of data entry.^2^ With the recent development of digital technology and the widespread use of mobile devices, DCT has overcome the shortcomings of traditional paper-based surveys and made it possible to conduct surveys more quickly. In recent years, DCT has been associated with effective control of Ebola hemorrhagic fever in Sierra Leone, tuberculosis in Botswana, and whooping cough in the United States.^3^

Ensuring that large numbers of people install DCT apps is essential for effective operation of DCT against COVID-19.^4-6^ Previous studies revealed that demographic, behavioral, psychological, and COVID-19-related factors all influence the installation of DCT apps.^1, 3, 7-9^ From the demographic aspect, ethnic minorities, migrants, low-income people, and elderly people were less likely to install the apps.^3^ From the behavioral aspect, non-smoking status was associated with high app acceptance, while lack of information technology skills and not having a smartphone were associated with low app acceptance.^1, 8^ From the psychological aspect, low app acceptance was associated with doubts about the effectiveness of DCT apps, anxiety about the collection of personal information, and distrust of the government and app developers.^8, 9^ From the COVID-19-related aspect, acceptance of new lifestyles under the COVID-19 epidemic such as use of hand sanitizers, avoidance of public transportation, and wearing masks was associated with app use.^7, 8^

Because the working-age population is at the epicenter of COVID-19 infections, it is important to increase the acceptance of DCT apps in this population.^10^ In other words, the public health benefits of adopting these apps need to be particularly recognized by the working-age population.^9, 11^ In countries where installation of DCT apps is optional, such as North America, Europe, Oceania, and Japan, the installation rates tend to be lower compared with countries where it is mandatory.^1, 7, 12^ However, few studies have evaluated the factors associated with acceptance of DCT apps using sufficient sample sizes of workers.^13^ The purpose of the present study was to evaluate the factors associated with acceptance of a DCT app for COVID-19 in the working-age population of Japan, where app installation is optional. The findings provide insights that can help to facilitate the adoption of DCT apps among workers.

## Methods

### Study design and participants

The present study had a cross-sectional design and used baseline questionnaire data from a prospective cohort research project in Japan, the Collaborative Online Research on the Novel-coronavirus and Work (CORoNaWork), between 22 and 26 December 2020.^14^ The study supplements another study from the CORoNaWork project that evaluated occupational factors associated with DCT adoption.^10^ During the research period, COVID-19 vaccines had not yet been introduced in Japan and the third wave of infection was at its peak, with more than 3,000 new infections per day.^15^ The inclusion criteria for the study were: full-time worker status, age between 20 and 65 years, and registration with an internet survey company. Invitations were sent by e-mail to 605,381 people registered with an internet survey company, of whom 55,045 people accessed the linked website. Of these, 33,302 participants who met the inclusion criteria provided informed consent and completed the baseline survey in an online format. After excluding 6,266 participants who gave incorrect answers to questions designed to identify inappropriate responses, 27,036 participants were eligible for the analysis.

### The Contact-Confirming Application (COCOA) DCT app in Japan

The COCOA is the only DCT app in Japan and was released by the Ministry of Health, Labour and Welfare in June 2020. The COCOA uses a decentralized approach through Bluetooth and is designed not to store personal information on a central server controlled by the government or the developer. Furthermore, downloading and deleting the COCOA is optional. If a person comes into contact with a COVID-19-positive person within a 1-m radius for more than 15 minutes, the person is notified as a close contact.^6^

### Dependent variable

The dependent variable in the study was the following question about adoption of the COCOA: “Have you downloaded the Contact-Confirming Application (COCOA)?” The response options were “yes” and “no”.

### Independent variables

Based on relevant studies, the independent variables were divided into four aspects: demographic, behavioral, psychological, and COVID-19 transmission-related variables.^1, 3, 7-9^ Demographic variables included sex, age, marital status, education, annual household income (in Japanese yen), occupation (desk work, work with interpersonal communication, manual work), and family members living together. Behavioral variables included smoking status, alcohol consumption, and regular doctor’s visits for chronic diseases. Psychological variables included fear of COVID-19 transmission. Variables related to COVID-19 transmission included wearing a mask, using hand sanitizer, willingness to be vaccinated against COVID-19, and presence of an acquaintance infected with COVID-19.

### Statistical analysis

Factors associated with COCOA adoption were evaluated by univariate and multivariate logistic regression analyses, and the results were presented as odds ratios [ORs] and 95% confidence intervals [CIs]. The multivariate analyses were adjusted for sex, age, marital status, education, and annual household income, based on relevant studies.^1, 3, 7-9^ The *P*-values were two-sided, and statistical significance was set at *P* < 0.05. All statistical analyses were performed using Stata/SE 16.1 software (StataCorp, College Station, TX, USA).

## Results

Table 1 shows the general characteristics of the study participants. Of the 27,036 participants, around half were men (51.1%), married (55.6%), educated at university or graduate school (48.7%), and engaged in desk work (49.8%). Regarding acceptance of the DCT app for COVID-19, 6,786 respondents (25.1%) reported that they had downloaded the COCOA.

**Table 1.** General characteristics of the study participants

|  | Total<br>N=27,036<br>n (%) | Acceptance of digital contact tracing<br>application for COVID-19 |  |
| --- | --- | --- | --- |
|  |  | Yes<br>n=6,786<br>n (%) | No<br>n=20,250<br>n (%) |
| Sex |  |  |  |
| Women | 13,222 (48.9) | 3,181 (46.9) | 10,041 (49.6) |
| Men | 13,814 (51.1) | 3,605 (53.1) | 10,209 (50.4) |
| Age |  |  |  |
| 20–39 years | 6,763 (25.0) | 1,669 (24.6) | 50,94 (25.2) |
| 40–49 years | 8,011 (29.6) | 1,886 (27.8) | 6,125 (30.2) |
| 50–59 years | 9,012 (33.4) | 2,359 (34.8) | 6,653 (23.9) |
| 60–65 years | 3,250 (12.0) | 872 (12.8) | 2,378 (11.7) |
| Marital status |  |  |  |
| Single | 9,164 (33.9) | 1,991 (29.4) | 7,173 (35.4) |
| Divorced or widowed | 2,843 (10.5) | 722 (10.6) | 2,121 (10.5) |
| Married | 15,029 (55.6) | 4,073 (60.0) | 10,956 (54.1) |
| Education |  |  |  |
| Junior high or high school | 7,321 (27.1) | 1,647 (24.3) | 5,674 (28.0) |
| Vocational school or college | 6,544 (24.2) | 1,552 (22.9) | 4,992 (24.7) |
| University or graduate school | 13,171 (48.7) | 3,587 (52.8) | 9,584 (47.3) |
| Annual household income |  |  |  |
| < 2,000,000 yen | 1,709 (6.3) | 277 (4.1) | 1,432 (7.1) |
| 2,000,000–3,999,999 yen | 5,548 (20.5) | 1,188 (17.5) | 4,360 (21.5) |
| 4,000,000–7,999,999 yen | 11,927 (44.1) | 2,925 (43.1) | 9,002 (44.5) |
| ≥ 8,000,000 yen | 7,852 (29.1) | 2,396 (35.3) | 5,456 (26.9) |
| Job type |  |  |  |
| Manual work | 6,641 (24.6) | 1,283 (18.9) | 5,358 (26.5) |
| Desk work | 13,468 (49.8) | 3,736 (55.1) | 9,732 (48.1) |
| Work involving interpersonal communication | 6,927 (25.6) | 1,767 (26.0) | 5,160 (25.5) |
| Family living together |  |  |  |
| No | 5,807 (21.5) | 1,400 (20.6) | 4,407 (21.8) |
| Yes | 21,229 (78.5) | 5,386 (79.4) | 15,843 (78.2) |
| Smoking |  |  |  |
| Never | 14,587 (54.0) | 3,495 (51.5) | 11,092 (54.8) |
| Past | 5,445 (20.1) | 1,551 (22.9) | 3,894 (19.2) |
| Current | 7,004 (25.9) | 1,740 (25.6) | 5,264 (26.0) |

Table 1. General characteristics of the study participants (cont.)
|  | Total | Acceptance of digital contact tracing application for COVID-19 |  |
| --- | --- | --- | --- |
|  |  | Yes | No |
|  | N=27,036 | n=6,786 | n=20,250 |
|  | n (%) | n (%) | n (%) |
| Alcohol intake |  |  |  |
| Never | 11,472 (42.4) | 2,699 (39.8) | 8,773 (43.3) |
| Occasionally | 4,547 (16.8) | 1,186 (17.5) | 3,361 (16.6) |
| Regular | 11,017 (40.8) | 2,901 (42.7) | 8,116 (40.1) |
| Regular doctor's visits for chronic diseases |  |  |  |
| No | 17,526 (64.8) | 4,008 (59.1) | 13,518 (66.8) |
| Yes | 9,510 (35.2) | 2,778 (40.9) | 6,732 (33.2) |
| Fear of COVID-19 transmission |  |  |  |
| No | 7,019 (26.0) | 1,049 (15.5) | 5,970 (29.5) |
| Yes | 20,017 (74.0) | 5,737 (84.5) | 14,280 (70.5) |
| Wearing a mask |  |  |  |
| Rarely/never | 682 (2.5) | 84 (1.2) | 598 (3.0) |
| Often | 3,046 (11.3) | 570 (8.4) | 2,476 (12.2) |
| Always | 23,308 (86.2) | 6,132 (90.4) | 17,176 (84.8) |
| Using hand sanitizer |  |  |  |
| Rarely/never | 3,740 (13.8) | 633 (9.3) | 3,107 (15.3) |
| Often | 8,282 (30.6) | 1,965 (29.0) | 6,317 (31.2) |
| Always | 15,014 (55.4) | 4,188 (61.7) | 10,826 (53.5) |
| Willing to receive the COVID-19 vaccine |  |  |  |
| No | 16,889 (62.5) | 3,467 (51.1) | 13,422 (66.3) |
| Yes | 10,147 (37.5) | 3,319 (48.9) | 6,828 (33.7) |
| Presence of an acquaintance infected with COVID-19 |  |  |  |
| No | 24,760 (91.6) | 5,947 (87.6) | 18,813 (92.9) |
| Yes | 2,276 (8.4) | 839 (12.4) | 1,437 (7.1) |

COVID-19: coronavirus disease 2019.

Table 2 presents the factors associated with acceptance of the DCT app for COVID-19. In the multivariate analyses, married (adjusted OR: 1.29; 95% CI: 1.17–1.43) and divorced/widowed (adjusted OR: 1.16; 95% CI: 1.09–1.25) individuals had significantly higher acceptance of the DCT app than single individuals. Similarly, people with a university or graduate school degree had higher acceptance of the DCT app than people with a junior high school or high school degree (adjusted OR: 1.19; 95% CI: 1.11–1.28). Higher annual income was associated with higher acceptance of the app (≥8,000,000 yen: adjusted OR: 2.08; 95% CI: 1.80–2.39). In contrast, acceptance of the app was lower among people living with family members (adjusted OR: 0.82; 95% CI: 0.76–0.89).

**Table 2.**
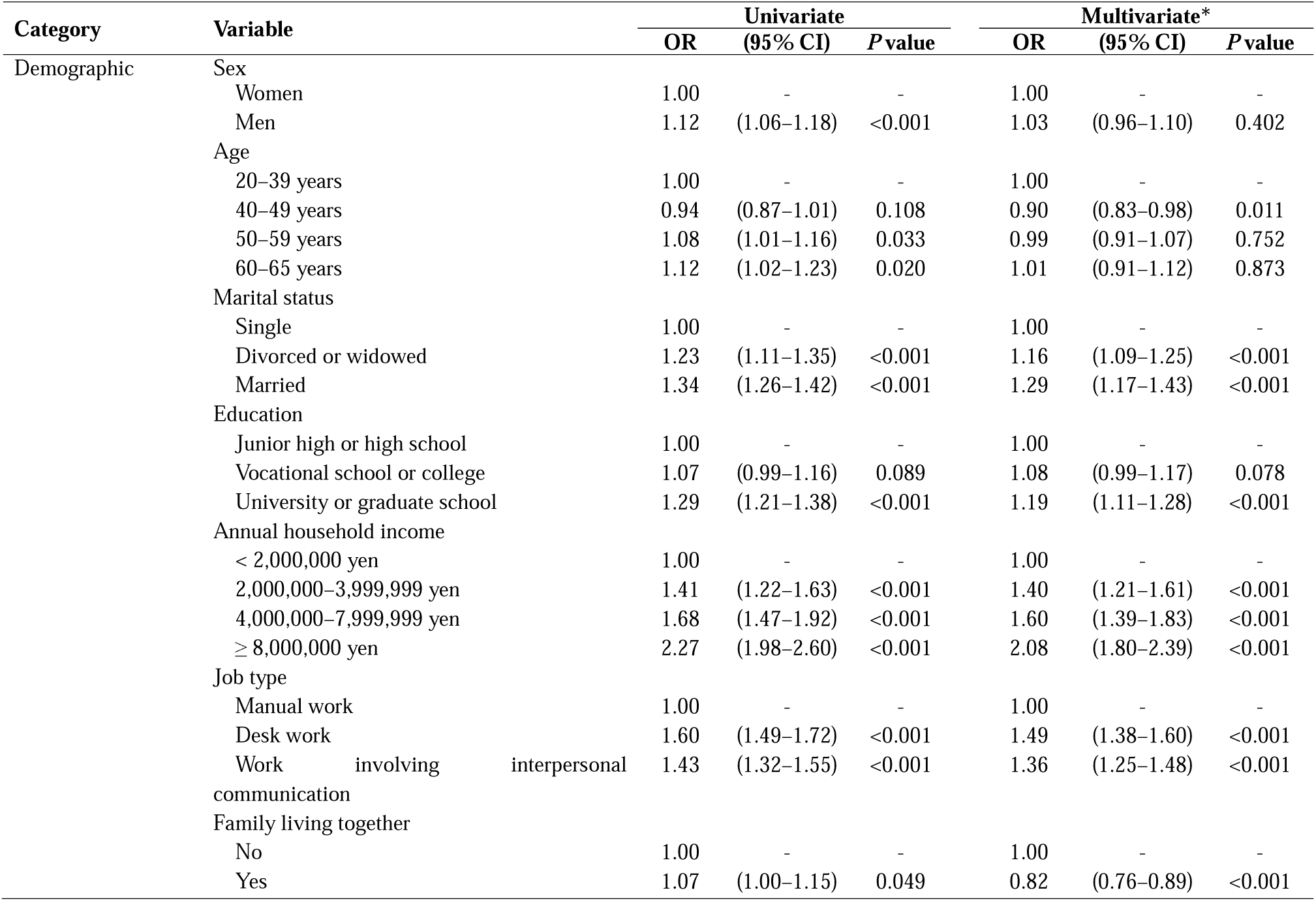
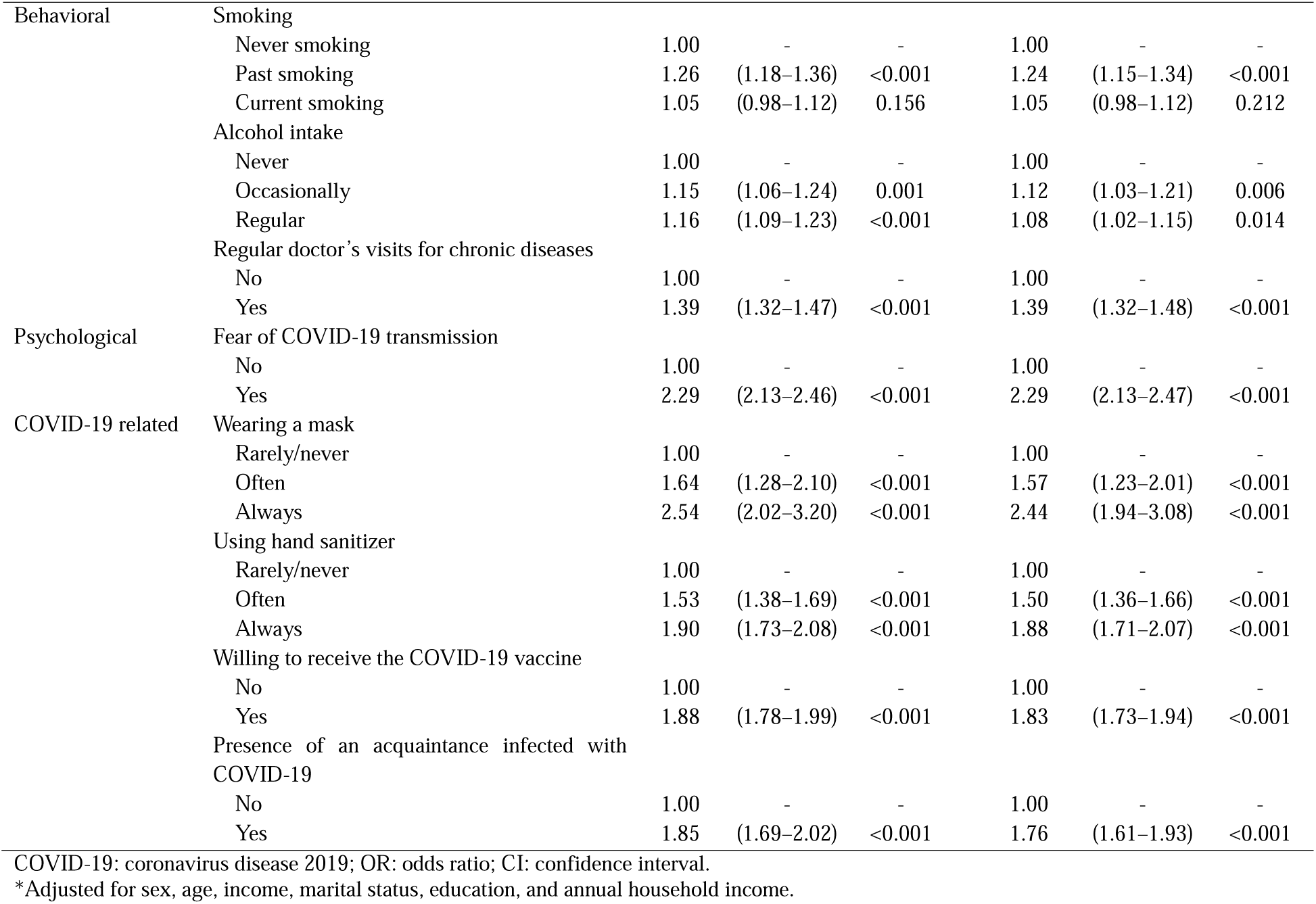
Factors associated with acceptance of the digital contact tracing application for COVID-19

From the behavioral aspect, past smokers were more accepting of the app than non-smokers (adjusted OR: 1.24; 95% CI: 1.15–1.34), but there was no significant difference for smokers. Patients who regularly visited the hospital for chronic diseases were more likely to accept the app than patients who did not (adjusted OR: 1.39; 95% CI: 1.32–1.48). From the psychological aspect, the odds of accepting the app were 2.29 times (95% CI: 2.13–2.47) higher in people who feared COVID-19 transmission. From the COVID-19-related aspect, people who always wore a mask (adjusted OR: 2.44; 95% CI: 1.94–3.08) or used hand sanitizer (adjusted OR: 1.88; 95% CI: 1.71–2.07) were more likely to accept the app than people who rarely or never performed these actions. Acceptance of the app had 1.83 times higher odds (95% CI: 1.73–1.94) among people who were willing to receive the COVID-19 vaccine than among people who were not. Similarly, people who had an acquaintance infected with COVID-19 had 1.76 times higher odds (95% CI: 1.61–1.93) of accepting the app than people who did not.

## Discussion

In the present study, the rate of downloading the COCOA in Japan was not very high at 25.1%. The results revealed that there were several factors associated with downloading of the DCT app for each of the demographic, behavioral, psychological, and COVID-19-related variables. Although different countries have differing policies toward DCT apps, the present results suggest that adoption of DCT apps may be influenced by individual characteristics. The findings of the present study have important implications for the widespread adoption of DCT apps in working-age populations in Japan and elsewhere.

From the demographic aspect, the present study showed that the DCT app was more likely to be accepted by people with married status, university graduation or above, higher income, and desk work. These findings suggest that relatively wealthy people in society are more likely to use DCT apps, consistent with previous studies conducted in many countries including the United States, the United Kingdom, Switzerland, and Japan.^8, 16-18^ In contrast, the present study found that living with family members tended to reduce the use of the DCT app, which was inconsistent with a previous study.^19^ Although the reason for the discrepancy remains unclear, it is possible that family interactions reduced anxiety about COVID-19, which in turn reduced the need for the DCT app. Conflicting results were also reported in another study. Specifically, Tomczyk et al.^20^ pointed out that greater perceived usefulness of DCT apps was associated with greater fear of COVID-19 infection and lower adoption of the apps. Therefore, people who are afraid of letting their families know that they are infected with COVID-19 may tend to avoid DCT apps.

From the behavioral aspect, individuals with regular doctor’s visits for chronic diseases and past smokers were more willing to adopt the DCT app, while current smokers showed no association in the present study. The severity of COVID-19 was reported to be higher in people with regular doctor’s visits for chronic diseases such as hypertension and respiratory disease, and in smokers.^21^ Therefore, the present results suggest that people with a greater risk of severe COVID-19 are more positive about adopting DCT apps. On the contrary, current smokers generally have limited access to health information, which may have attenuated the result.^22^ Measures are needed to facilitate adoption of DCT apps by these people. For example, it may be useful for family physicians to encourage their patients to adopt DCT apps.

From the psychological aspect, fear of COVID-19 transmission was strongly associated with higher acceptance of the DCT app, consistent with a previous study in the United Kingdom that found a positive association between risk perception for COVID-19 infection and acceptance of DCT apps.^17^ Kawakami et al.^19^ found that installation of a DCT app mitigated the psychological distress associated with the risk of COVID-19 infection. Taken together, the findings suggest that fear of COVID-19 transmission increases the adoption of DCT apps and that appropriate adoption of DCT apps may reduce anxiety about COVID-19.

From the COVID-19-related aspect, wearing a mask, using hand disinfection, willingness to be vaccinated against COVID-19, and presence of an acquaintance infected with COVID-19 led to greater likelihood of adopting the DCT app. A previous study in Singapore demonstrated similar findings for mask wearing and hand sanitizer use.^7^ A similar trend was also observed in a study conducted in the United States, wherein people infected with COVID-19 had a higher acceptance rate of DCT apps.^18^ Based on these findings, posting Quick Response codes for DCT apps on the packages of masks and hand sanitizers and at vaccination sites may be an effective measure. A function for inviting friends within the apps may also be effective.^9^

There are several limitations to this study. First, we recruited participants who were registered with an internet research company. Therefore, we may have selected people who actively used the internet, smartphones, and apps. However, we consider this potential selection bias to be negligible because the COCOA downloading rate announced by the Ministry of Health, Labour and Welfare in December 2020 was 20.8%,^23^ and thus close to the rate in the present study (25.1%). Second, the present study used cross-sectional data for a single period of time. Therefore, the findings can only suggest associations between factors for this given period of time, and cannot capture changes over time within individuals. Third, in the multivariate logistic regression analyses, there was a possibility of unmeasured confounding. Unmeasured personal habits and behavioral patterns may have influenced the adoption of the DCT app.

## Conclusions

The present study revealed factors associated with adoption of a DCT app among the working-age population in Japan. Use of the COCOA among Japanese workers was not sufficient, and it is important to develop an approach based on the present findings. We found that relatively wealthy people in society were more likely to use the DCT app. In addition, people with a greater risk of severe COVID-19 were more positive about adopting the DCT app. Fear of COVID-19 transmission increased adoption of the DCT app, suggesting that appropriate adoption of DCT apps can reduce anxiety about COVID-19. Because wearing a mask, using hand disinfection, and willingness to be vaccinated against COVID-19 were associated with greater likelihood of adopting the DCT app, posting Quick Response codes for DCT apps on the packages of masks and hand sanitizers and at vaccination sites may be an effective measure.

## Data Availability

No data are available

## Acknowledgments

The current members of the CORoNaWork Project, in alphabetical order, are: Prof. Yoshihisa Fujino (present chairperson of the study group), Dr. Hajime Ando, Prof. Hisashi Eguchi, Dr. Kazunori Ikegami, Dr. Arisa Harada, Dr. Ayako Hino, Dr. Tomohiro Ishimaru, Dr. Kyoko Kitagawa, Ms. Ning Liu, Dr. Kosuke Mafune, Prof. Shinya Matsuda, Dr. Ryutaro Matsugaki, Prof. Koji Mori, Dr. Keiji Muramatsu, Dr. Masako Nagata, Dr. Tomohisa Nagata, Prof. Akira Ogami, Dr. Rie Tanaka, Dr. Seiishiro Tateishi, Dr. Kei Tokutsu, and Prof. Mayumi Tsuji. All members are affiliated with the University of Occupational and Environmental Health, Japan.

